# Avidity as a marker of protection against hepatitis E during a genotype 1 outbreak in South Sudan

**DOI:** 10.64898/2026.09.03.26362164

**Authors:** Camille Beatrice Gaza Bionda Valera, Catia Alvarez, Amy Dighe, Iza Ciglenecki, Isabella Eckerle, Robin C Nesbitt, Yoann Sarmiento, Etienne Gignoux, John Rumunu, Vincent Kinya Asilaza, Benjamin Meyer, Andrew S Azman

**Affiliations:** Institute of Global Health, University of Geneva, Geneva, Switzerland; Geneva Center for Emerging Viral Diseases, University Hospitals of Geneva, Geneva, Switzerland; Department of Epidemiology, Johns Hopkins Bloomberg School of Public Health, Baltimore, Maryland, USA; Department of Infectious Disease Epidemiology, Imperial College London, London, United Kingdom; Médecins Sans Frontières, Geneva, Switzerland; Epicentre, Paris, France; South Sudan Ministry of Health, Juba, South Sudan; Center of Vaccinology, Department of Pathology and Immunology, University of Geneva, Geneva, Switzerland

**Author notes:** Corresponding Author: Andrew S. Azman, PhD, Institute of Global Health, University of Geneva, 9 Chemin des Mines 1202 Geneva, Switzerland.

**Keywords:** hepatitis E, hepatitis E virus (HEV), avidity, anti-HEV IgG avidity

## Abstract

**Background:** In settings where hepatitis E virus (HEV) is highly endemic, anti-HEV IgG seroprevalence approaches saturation, yet how often disease occurs in previously infected individuals, and how protective natural genotype 1 immunity is, remains unmeasured. IgG concentration cannot answer this as levels rise within days of symptom onset masking any pre-existing levels; IgG avidity, maturing over months, may serve as a useful tool to understand reinfections.

**Methods:** We enrolled 1007 suspected HEV patients and measured anti-HEV IgG avidity in IgG-positive confirmed cases and an age-stratified sample of IgG-positive test-negatives (n = 366) in enhanced surveillance around a 2022 vaccination campaign in Bentiu, South Sudan. Using high avidity (avidity index ≥50%) as a marker of past infection, we estimated (i) the reinfection fraction, (ii) protection from prior infection using a test negative design and (iii) associations with viral load and infection severity.

**Results:** Median avidity index was 11% in cases versus 77% in test-negatives. Among unvaccinated cases, 11.6% (28/242) carried high-avidity IgG (indicative of mature immunity and therefore reinfection), rising with age (odds ratio 1.6 per decade, p=0.006). Mature immunity was associated with 95% lower odds of disease (adjusted OR 0.05, 95% CI 0.03–0.08), robust to threshold and case-definition. High-avidity cases were less often viremic (29% vs 78%) at matched time since onset, though we found no differences in liver function biomarkers between patients with high-avidity and low-avidity.

**Conclusions:** Most HEV disease occurred in previously uninfected individuals, and prior immunity was strongly protective against detected disease. Avidity provides a population-level tool to read the immune landscape where seroprevalence may be uninformative.

**Short Summary:** Using IgG avidity to distinguish between past from recent hepatitis E infection in a highly endemic setting, we found most disease occurred in previously uninfected individuals. Prior infection was associated with 95% lower odds of disease, demonstrating strong natural immunity.

## Introduction

Hepatitis E virus (HEV) is a leading cause of acute viral hepatitis globally [1–3]. Genotypes 1 and 2 predominate across Asia and Africa, where they sustain endemic transmission and periodically cause large-scale outbreaks, often in populations with poor access to safe water [4,5]. Genotype 1 infections are particularly significant as 1 in 4 infected pregnant women die from fulminant hepatic failure [6]. A three-dose recombinant vaccine with high efficacy, Hecolin (Innovax, Beijing, China), is licensed in several countries and available for use [7–9].

The extent to which prior HEV infection protects against subsequent infection and disease is a key parameter that shapes transmission and can inform disease control and prevention strategies. In a cohort study in China, baseline IgG seropositivity, a proxy for previous infection, was associated with a 58.5% (95% CI: 44.8-68.7%) reduced risk of subsequent genotype 4 HEV reinfection and a 71.5% (95% CI: −37.9-94.1%) reduced risk of subsequent hepatitis E disease, although the latter estimate was highly uncertain [10]. Comparable estimates do not exist for other genotypes, including genotype 1, which causes substantial disease in settings where repeated HEV exposure is common.

Estimating protection from prior infection ordinarily requires prospective measurement of immune status before subsequent exposure or disease, which is difficult to achieve outside intensively followed cohorts. This challenge is particularly acute in highly endemic settings where repeated exposure drives anti-HEV IgG seropositivity to high, sometimes near-universal, levels, reflecting a mix of recent and more distal infections [4,6,11]. Moreover, IgG develops rapidly during acute HEV infections. In a serial follow-up of genotype 3 infections, IgG antibodies at uniformly low-avidity were detected in essentially all patients at acute presentation [12], and in frequently sampled viremic blood donors, IgM and IgG positivity rose in parallel [13]. Equivalent serial data do not exist for genotype 1, but the near-universal IgG positivity among patients with an acute genotype 1 infection sampled within days of onset is consistent with comparable kinetics [8,11]. Thus, among patients sampled after disease onset, IgG concentration alone cannot distinguish antibodies that predated the current infection from those elicited by it.

IgG avidity provides the timescale that concentration lacks. Avidity, the aggregate binding strength of the antibody-antigen interaction, matures over months as the B cell response undergoes affinity maturation, with low-avidity antibodies predominating during primary infection and progressively replaced by high-avidity IgG [12,14–16]. Since secondary immune responses are largely memory-derived and of high avidity, a current infection does not lower avidity. The avidity index has been used as the basis for distinguishing primary from repeat infection across dengue, measles, rubella, cytomegalovirus and toxoplasmosis as well as HEV [15,17–21]. Evidence also shows that avidity may be a more specific indicator of antibody-mediated protection than concentration as long-term follow-up of the Hecolin efficacy trial found efficacy persisting for a decade while IgG concentrations fell ~90-fold [7,10,22,23], and in experimentally reinfected rhesus macaques, high baseline avidity was protective while lower avidity resulted in attenuated disease with reduced viremia [24]. Despite this evidence, avidity has not been measured at scale in a genotype 1 outbreak, where prior exposure is widespread and its interpretive value would be greatest.

The Bentiu displaced persons camp in South Sudan hosts approximately 100,000 residents and has experienced protracted HEV transmission, with anti-HEV IgG seropositivity exceeding 90% among patients presenting with acute jaundice [8]. Hecolin was reactively deployed for the first time during the 2022 HEV outbreak, alongside an enhanced surveillance platform that enrolled patients with suspected HEV and collected paired serum samples [8]. Leveraging these samples, we measured anti-HEV IgG avidity to address three questions: (i) what proportion of detected genotype 1 disease occurs in individuals with evidence of pre-existing mature immunity (e.g. high avidity); (ii) whether pre-existing mature immunity is associated with reduced odds of confirmed HEV disease; and (iii) whether pre-existing mature immunity is associated with viral replication and biochemical markers of liver injury.

## Methods

### Setting, surveillance, and case definitions

Between May and December 2022, enhanced surveillance in the Bentiu camp enrolled patients meeting the suspected HEV case definition of acute jaundice syndrome. Serum was collected at presentation, with a follow-up visit requested at least two weeks later. We tested all participants for anti-HEV IgM and IgG by ELISA (Wantai) and for HEV RNA by PCR (Mikrogen Diagnostik). A confirmed case was positive for anti-HEV IgM and/or HEV RNA; a test-negative participant was negative for both. We recorded time from symptom onset to presentation, sex, age, and vaccination history (number of effective doses, counted from 14 days after administration). Participants with ambiguous test results, 2 IgG-indeterminate test-negatives, were excluded from analyses requiring an immune state.

### Laboratory testing

A series of liver function tests (alanine aminotransferase [ALT] and aspartate aminotransferase [AST]) using a Reflotron or SimplexTAS machine were conducted in the hospital laboratory. All specimens were prepared for storage and transport by separating plasma from whole blood by centrifugation and frozen at −20 °C within 6 hours of collection. Samples were transported to the Geneva Center for Emerging Viral Diseases at the Geneva University Hospitals, Switzerland on dry ice with temperature loggers to ensure a −80 °C cold chain. Upon arrival at the reference laboratory, samples were stored at −80 °C until testing.

RT-PCR, IgM ELISA, IgG ELISA, and avidity testing were conducted at the University of Geneva. Prior to RT-PCR tests, we extracted RNA from plasma using the NucliSens easyMAG instrument (BioMérieux, Marcy-l’Étoile, France). We then used the Mikrogen Diagnostik ampliCube HEV 2.0 Quant real-time quantitative PCR (RT-PCR) system according to the manufacturer’s instructions to detect HEV RNA with primers and probes specific for genotypes 1, 2, 3, and 4. We used a cycle threshold positivity cutoff value of ≤42. To detect IgM and IgG antibodies in plasma samples, we used Wantai HEV-IgM ELISA (WE-7196) and Wantai HEV-IgG ELISA (WE-7296) according to the manufacturer’s instructions (Wantai BioPharm, Beijing, China) on an automated ELISA system.

For avidity testing, we modified the Wantai HEV-IgG ELISA assay to include a urea wash step. We measured avidity in all IgG-positive confirmed cases with a sufficient volume of leftover serum remaining after primary confirmatory testing, and in an age-stratified sample of IgG-positive test-negative participants (Supplementary Figure S1). Each plasma sample was tested in duplicate under both standard and urea-treated conditions on the same plate. Following the initial antigen-antibody incubation, half of the plate wells were washed according to the standard kit protocol, while the remaining wells were washed with 5M urea prepared in the kit’s diluted washing buffer. Plates were washed five times under both conditions, and the assay was then continued following the standard ELISA procedure. Optical density (OD) values were measured at 450/600 nm using a microplate reader. We calculated the avidity index (AI) as the ratio of the OD value obtained in the urea-treated wells to that of the corresponding control wells for each sample, expressed as a percentage.

To ensure accurate and reproducible measurements, we diluted samples prior to testing to obtain OD values within the linear range of the ELISA (0.5-2.7) in line with a previously published approach [12]. Dilution factors varied between samples, ranging from 1:5 to 1:16,500. A standard, prepared as a pooled sample from ten individual plasma, was included on each plate to enable comparison across assay runs and to control for inter-plate variability. This approach minimized signal saturation or loss and ensured reliable calculation of the avidity index under both standard and urea-treated conditions. To ensure that urea treatment did not affect conformation or binding of antigen on the plate, we incubated the plate either with regular wash buffer or with 5M urea before performing the ELISA with positive and negative control samples as well as several dilutions of the serum standard. ODs in treated and untreated wells remained highly similar indicating no effect of urea treatment on antigen integrity (Supplementary Figure S2).

We also assessed the presence of IgG antibodies against the ORF2 Wantai antigen using a bead-based Luminex immunoassay. Magnetic microspheres were covalently coupled to the ORF2 antigen following the manufacturer’s instructions (MagPlex®-C Microspheres, MC10013). Briefly, beads were activated and incubated with the antigen at a concentration of 5 µg per million beads under optimized coupling conditions. Excess antigen was removed through washing steps and beads were blocked using PBS with 1% BSA. The coupled beads were then resuspended in an assay buffer and dispensed into a 96-well plate. Serum samples from Bentiu were diluted 1:1000 and tested in duplicate. Diluted samples were incubated with antigen-coated beads to allow antibody binding. Following incubation, unbound components were removed via washing. Bound antibodies were detected using a phycoerythrin (PE)-conjugated mouse anti-human IgG Fc secondary antibody (NB110-8347PE, Novus Biologicals) at a 1:1000 dilution. After a final wash, beads were resuspended in sheath fluid and analyzed on a Luminex xMAP INTELLIFLEX instrument. Signals were recorded as median fluorescence intensity (MFI). Each assay plate included a standard for calibration, as well as positive and negative controls to monitor sensitivity and background signal, ensuring consistent assay performance across runs.

### Statistical analysis

The exposure of interest was prior HEV infection, marked by high avidity IgG. As there is no universal cutoff for “high” avidity, we defined high as having an avidity index of ≥50% for the main analyses and explored alternative thresholds in sensitivity analysis. We used high avidity as a measure in three primary analyses estimating the reinfection fraction (ii) estimating protection associated with high avidity and (iii) the association between high avidity and markers of disease severity.

We estimated the reinfection fraction as the proportion of avidity-tested confirmed cases with high-avidity IgG. This analysis was restricted to unvaccinated cases, in whom high avidity IgG can develop only from prior infection, not vaccination. We used logistic regression with age as a continuous variable to quantify how this fraction varied with age.

In our analysis of protection, we derived the estimand and adjustment set from an explicit causal diagram (Supplementary Figure S3; d-separation verified in dagitty, Supplementary Table S1), in which age was the only measured common cause of prior and current infection. The analytic population for the protection models comprised all unvaccinated participants with an observed exposure (IgG-negative, or IgG-positive with a measured AI) presenting within 30 days of symptom onset (Supplementary Figure S1). Since sampling of test-negative participants was dependent on IgG status, we applied inverse-probability sampling weights to the subsampled IgG-positive test-negative participants, each the ratio of the number eligible to the number selected within that participant’s age band (580 eligible, 99 selected overall). The resulting band weights were 4.0, 3.2, 3.1, 16.0, 3.6, and all other participants were assigned a weight of 1.

We estimated protection, defined as 1 - adjusted odds ratio, using weighted logistic regression (survey design; svyglm) of confirmed-case status on high-avidity IgG, adjusted for age (spline), sex, and time since symptom onset, among unvaccinated participants presenting within 30 days. For sensitivity analyses, we fit unweighted and linear-age models, and a model restricting cases to PCR-confirmed disease. All 95% CIs are design-based Wald intervals, and we assessed robustness to the avidity threshold by refitting models across AI cutoffs from 15% to 90%.Given that prior immunity may accelerate viral clearance, we treated viremia (PCR positivity) at presentation as the primary outcome in analyses related to severity, modeling it as a function of avidity adjusting for time since onset (and, secondarily, age and sex). Among viremic (PCR-positive) cases, we compared viral load, ALT, and total bilirubin by avidity, and we report overall (IgM-or-PCR) comparisons as sensitivity analyses. Follow-up sampling times varied widely, so we analyzed avidity maturation as a function of time since onset using a linear mixed model with a per-patient random intercept.

Analyses used R version 4.3 (survey, splines, sandwich/lmtest, logistf, lme4, dagitty). Data and code to reproduce analyses are available at https://github.com/GenevaIDD/hev-igg-avidity.

### Ethics

The parent study was approved by the MSF Ethics Review Board (ERB #2167) and the South Sudan Ministry of Health Research Ethics Board (RERB-MOH #57/27/09/2022). All adults provided written informed consent to participate, and individuals younger than 18 years provided written informed assent alongside written informed consent from their guardians. All study participants provided consent to subsequent use of their samples for further HEV-related research.

## Results

A total of 1007 individuals meeting the suspected HEV case definition were enrolled between May and December 2022: 276 confirmed cases (HEV RNA and/or anti-HEV IgM positive) and 731 test-negative participants (Table 1). At the initial visit, 84.5% (n=851) were anti-HEV IgG positive and 15.3% (n=154) negative; 0.2% (n=2) had indeterminate results. We measured avidity for 366 individuals with available blood samples (267 confirmed cases, 99 test-negative participants; Figure 1A, Supplementary Table 2). Analyses of protection associated with high avidity antibodies drew on the 412 participants with an observed exposure (221 confirmed cases, 191 test-negative participants), all unvaccinated and presenting within 30 days of onset (Supplementary Figure S1).

**Table 1.** Characteristics of participants enrolled in enhanced HEV surveillance, Bentiu IDP camp, South Sudan, May–December 2022.

|  | Confirmed HEV case<br>(N=276) | Test-negative<br>(N=731) | Overall<br>(N=1007) | p† |
| --- | --- | --- | --- | --- |
| **Age, years, median [IQR]** | 9.5 [4.8, 17.7] | 17.5 [6.3, 27.9] | 14.5 [5.5, 25.2] | <0.001 |
| **Age group, n (%)** |  |  |  | <0.001 |
| 0–5 years | 75 (27.2) | 156 (21.3) | 231 (22.9) |  |
| 6–10 years | 67 (24.3) | 87 (11.9) | 154 (15.3) |  |
| 11–15 years | 46 (16.7) | 80 (10.9) | 126 (12.5) |  |
| 16–39 years | 80 (29.0) | 330 (45.1) | 410 (40.7) |  |
| 40+ years | 8 (2.9) | 78 (10.7) | 86 (8.5) |  |
| **Sex, n (%)** |  |  |  | 1.000 |
| Female | 127 (46.0) | 336 (46.0) | 463 (46.0) |  |
| Male | 149 (54.0) | 395 (54.0) | 544 (54.0) |  |
| **Days from symptom onset, n (%)** |  |  |  | 0.001 |
| 0–5 days | 142 (51.4) | 350 (47.9) | 492 (48.9) |  |
| 6–30 days | 107 (38.8) | 252 (34.5) | 359 (35.7) |  |
| 31–100 days | 23 (8.3) | 79 (10.8) | 102 (10.1) |  |
| Missing | 4 (1.4) | 50 (6.8) | 54 (5.4) |  |
| **Effective vaccine doses, n (%)** |  |  |  | <0.001 |
| 0 | 250 (90.6) | 552 (75.5) | 802 (79.6) |  |
| 1 | 17 (6.2) | 58 (7.9) | 75 (7.4) |  |
| ≥2 | 9 (3.3) | 121 (16.6) | 130 (12.9) |  |
| **Case ascertainment, n (%)** |  |  |  | — |
| HEV RNA positive | 194 (70.3) | 0 (0.0) | 194 (19.3) |  |
| Anti-HEV IgM positive | 262 (94.9) | 0 (0.0) | 262 (26.0) |  |
| **Baseline anti-HEV IgG, n (%)** |  |  |  | <0.001 |
| Positive | 271 (98.2) | 580 (79.3) | 851 (84.5) |  |
| Negative | 5 (1.8) | 149 (20.4) | 154 (15.3) |  |
| Indeterminate | 0 (0.0) | 2 (0.3) | 2 (0.2) |  |
| **Baseline anti-HEV IgG avidity index, %, median [IQR]** | 11.2 [5.8, 28.2] | 77.4 [66.6, 87.4] | 20.0 [7.7, 69.2] | <0.001 |
| **ALT, U/L, median [IQR]** | 164.5 [15.0, 816.2] | 15.8 [11.8, 24.6] | 17.8 [12.1, 41.8] | <0.001 |
| Missing, n (%) | 68 (24.6) | 232 (31.7) | 300 (29.8) |  |
| **Total bilirubin, µmol/L, median [IQR]** | 3.88 [0.50, 10.60] | 0.50 [0.50, 0.72] | 0.50 [0.50, 1.41] | <0.001 |
| Missing, n (%) | 90 (32.6) | 237 (32.4) | 327 (32.5) |  |
| **Avidity measured, n (%)** | 267 (96.7) | 99 (13.5) | 366 (36.3) | — |

† Wilcoxon rank-sum for continuous variables, Fisher's exact for categorical, comparing confirmed cases with test-negative participants. Case-ascertainment and avidity-measured rows differ by definition/design and are not tested.
\* Normal ALT for males is up to 41 U/L (37°C), and up to 32 U/L (37°C) for females [25].
\*\* Normal total bilirubin range is 1.71 to 20.5 µmol/L [26].

**Figure 1.**
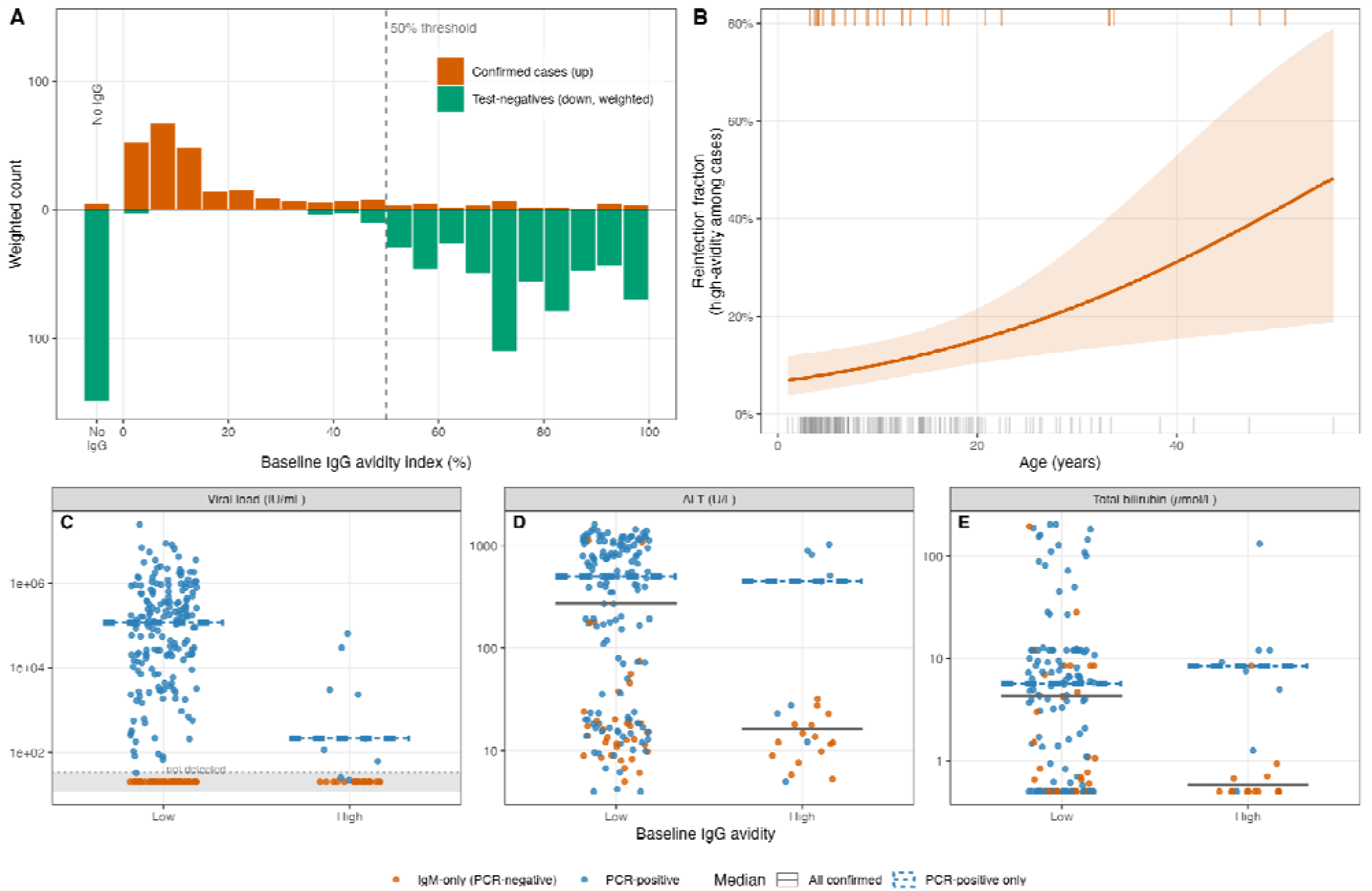
(A) Anti-HEV IgG avidity distribution - confirmed cases (up) vs. weighted test-negative participants (down), with the no-IgG block and 50% threshold. (B) Reinfection fraction (high-avidity IgG among confirmed cases) by age: fitted logistic curve with 95% CI; rug marks show individual high- (top) and low-avidity (bottom) cases. (C-E) Severity markers in confirmed cases by baseline IgG avidity (low vs. high). Points represent individuals colored by infection status, blue for PCR-positive vs. orange for IgM-only (PCR-negative), with horizontal lines showing group medians. In the viral load panel, viremic (PCR+) cases are shown at their viral load and aviremic (IgM-only) cases at a “not detected” floor; only the PCR-positive median is shown as viral load is undefined for aviremic cases.

Avidity differed strongly by case status. Confirmed cases had a median AI of 11% (IQR 6-28), with 13% (34/267) having high avidity (Table 1; Supplementary Table S2). Test-negative participants had a median AI of 77% (IQR 67-87), with 94% (93/99) having high avidity. Among test-negative participants, seroprevalence and mature immunity increased with age (Supplementary Figure S4). Among 242 unvaccinated confirmed cases, 11.6% (28/242) had high-avidity IgG, indicative of mature immunity and therefore reinfection (Supplementary Table S2). Since primary infections cannot generate high-avidity antibodies within days of onset, these represent true prior infections, though the estimate is likely higher, as some reinfections whose prior IgG had waned may have presented before their recalled high-avidity response is detected. The reinfection fraction increased with age (odds ratio 1.6 per decade, 95% CI 1.1– 2.2, p=0.006; Figure 1B), consistent with the accumulation of immunity over time (Supplementary Figure S5).

Relative to the absence of serological evidence of prior immunity, high-avidity IgG was associated with 95% lower odds of confirmed HEV disease (adjusted OR 0.05, 95% CI 0.03–0.08; weighted, spline-adjusted; Supplementary Table S3). The unweighted estimate was more conservative (81%), consistent with weighting correcting the undersampling of high-avidity test-negative participants. Protection was robust to the avidity threshold (82–95% across cutoffs from 15% to 90%; Supplementary Table S4) and when restricting cases to PCR-confirmed disease (98%; Supplementary Table S3).

High-avidity IgG identifies prior infection with high specificity but imperfect sensitivity (94%; 93/99 of IgG-positive test-negative controls). This sensitivity is unlikely to differ between cases and non-cases; given that a current infection does not lower current avidity, this misclassification is likely non-differential and should bias the estimate toward the null.

In evaluating the association between avidity and biomarkers of infection severity and viremia, we found that among confirmed cases, high-avidity individuals were less likely to be viremic than those with low-avidity (29% vs 78% PCR-positive; Supplementary Table S2). This did not reflect presentation as high-avidity IgM-only cases presented as early as viremic cases (median 5 days from onset in both), and after adjusting for time since onset, high avidity was associated with ~90% lower odds of viremia (adjusted OR 0.10, 95% CI 0.04–0.23; OR 0.06 with further adjustment for age and sex). We found no association between IgG levels and viral load (Supplementary Figure S6).

Confirmed cases with high avidity also had lower ALT and bilirubin across all confirmed cases, but this reflected case composition rather than an effect on the liver: high-avidity cases were predominantly aviremic (IgM-only), and among PCR-positive cases alone biochemical severity did not differ with ALT (median 449 vs 500 U/L, p=0.44) and total bilirubin (8.4 vs 5.7 µmol/L, p=0.44) being similar between high- and low-avidity cases (9–10 high-avidity cases; Figure 1C-E). Many high-avidity IgM-only cases may not represent acute hepatitis E; since anti-HEV IgM can persist for months, some likely reflect jaundice from another cause in a person with a past HEV infection. Consistent with this, high-avidity confirmed cases were about twice as likely as low-avidity cases to have a competing cause of jaundice (positive malaria RDT, HBsAg, or HAV/HBV PCR; 32% vs 18%; Supplementary Table S5) and excluding or reclassifying these cases did not materially change our primary estimates (Supplementary Table S6).

Prior immunity therefore appears to act predominantly as a barrier against establishing viremic infection rather than attenuating hepatic injury once viremia is present; the apparent ALT and bilirubin reductions across all confirmed cases reflect the enrichment of high-avidity cases for aviremic infection.

## Discussion

In this large-scale measurement of anti-HEV IgG avidity in a genotype 1 outbreak, most detected disease occurred in individuals with no evidence of prior infection. Among unvaccinated patients with confirmed HEV infection, only 11.6% had high-avidity IgG, indicative of prior infection. The same analysis yields, to our knowledge, the first field estimate of protection from natural genotype 1 immunity, at approximately 95%. These estimates are internally consistent. ~77% of test-negative participants had mature immunity (seropositive with high-avidity IgG, weighted for the age-stratified sampling; Supplementary Figure S4). Assuming this immune fraction reflects the wider population, then at 95% protection, the estimated reinfection would be ~13%, close to the observed 11.6%. In this setting, seroprevalence appears to broadly reflect the distribution of protective immunity, which is reassuring for serosurvey-based planning, though this relationship may not hold across settings, especially during acute outbreaks.

Prior immunity acted principally by preventing productive viremia. high-avidity cases were much less likely to be viremic at matched time since symptom onset, indicating rapid viral clearance rather than late presentation. Among the minority who remained viremic, liver injury markers were similar, suggesting that prior immunity blocks the establishment of replicating infection rather than modifying hepatocellular injury once replication is underway, in accordance with the largely immune-mediated pathogenesis of HEV liver injury.

We interpret avidity as a marker of protective immune status rather than as its cause. Whether avidity is mechanistically protective or a non-mechanistic correlate cannot be resolved from observational data, since avidity covaries with concentration, neutralizing activity, and cellular immunity [27]. Furthermore, the shape of the dose-response relationship is unable to discriminate a mechanistic effect from a non-mechanistic correlate; a mechanistically protective response could also show saturation above the threshold. This interpretation is standard in measles serology, where a low-avidity case in a vaccinated individual is called a primary vaccine failure, and a high-avidity case secondary vaccine failure or reinfection [18]. The same framework applies here to both vaccinated and naturally immune individuals.

Our study has several limitations; two notably bias the protection estimate and act in opposing directions. First, our measured outcome is detected HEV disease rather than infection, and the exposure (prior infection) lies on the causal path to detection. Given that prior immunity reduces the risk of becoming a detected case (PCR- or IgM-defined) by clearing virus (Supplementary Figure S3), reinfections that resolve subclinically never register as cases and instead fall into the test-negative group. This inflates the estimate by combining true protection with reduced detectability. Accordingly, our estimand should be interpreted as protection against detected clinical HEV disease rather than protection against infection. Second, and in the opposite direction, IgM can persist for many months [28], so some IgM-only cases may represent non-HEV jaundice rather than current infection. Restricting cases to PCR-confirmed disease raised the estimate to 98%, which indicates that misclassification of IgM-only cases does not drive our finding (Supplementary Table S6).

Other study limitations concern misclassification of previously infected individuals into the reference group. For instance, non-maturation of the antibody response places truly infected individuals below the avidity threshold, though this affects at most 6% of our study population, as shown by the control distribution (IgG-positive, test-negative participants with low-avidity). Seroreversion is another concern since it would make IgG-negativity an imperfect proxy for a truly naive immune state, as previously infected individuals whose IgG waned below detection fall into the reference group, which age adjustment cannot fully correct for. Reported annual seroreversion risk varies widely, from roughly 2% in rural Bangladesh and 5% in China, to 15% in a cohort in the Sitakunda sub-district of Chattogram, Bangladesh, where seroreversion rates were also found to be significantly faster among children [29–31]. Finally, several limitations concern generalization rather than internal validity. Avidity assays are research methods lacking an international reference reagent, with unstandardized per-sample dilutions and wash conditions across laboratories. Surveillance only captures acute jaundice presenting to care and is unable to assess subclinical reinfection. We also do not estimate vaccine effectiveness as the vaccinated stratum in our study was small (n = 42), and among those vaccinated, avidity largely reflected dose number (median AI = 10% after one dose vs 68% after two doses), it therefore carries no independent protection signal and is only reported descriptively.

In a genotype 1 HEV outbreak, our results demonstrate that prior natural infection is associated with substantial protection against detected clinical HEV disease, acting mainly by controlling viral replication, with most disease occurring among previously uninfected individuals. Anti-HEV IgG avidity provides a practical population-level tool for characterizing the immune landscape and estimating reinfection and protection, with potential value as a biomarker for vaccine and epidemiological studies.

## Data Availability

The code to reproduce analyses and a minimal dataset can be accessed at https://github.com/GenevaIDD/hev-igg-avidity.

## Acknowledgements

The parent study, through which all patients were recruited and samples collected, was funded by MSF. The additional lab and statistical analyses conducted in this manuscript were funded by the Gates Foundation (INV-064270 to ASA). The code to reproduce analyses and a minimal dataset can be accessed at https://github.com/GenevaIDD/hev-igg-avidity.

The authors would like to thank all participants in Bentiu and all the MSF field staff involved in patient recruitment and followup.

## Author Contributions

Conceptualization: BM, IC, ASA; Data curation: CBGBV, AD, RN, ASA; Formal Analysis: CBGBV, CA, AD, ASA; Funding acquisition: ASA; Investigation: CA, YS, VKA, BM, ASA; Methodology: BM, ASA; Project administration: BM, ASA, RN; Resources: BM, ASA; Software: ASA, CBGBV; Supervision: RN, IC, IE, BM, ASA; Validation: BM, ASA; Visualization: CBGBV, ASA; Writing – original draft: CBGBV, CA, ASA; Writing – review & editing: CBGBV, CA, AD, IC, IE, RCN, YS, EG, JR, VKA, BM, ASA

## Notes

### Competing Interest Statement

The authors have declared no competing interest.

### Author Declarations

The parent study was approved by the Medecins Sans Frontieres Ethics Review Board (ERB #2167) and the South Sudan Ministry of Health Research Ethics Board (RERB-MOH #57/27/09/2022).

