## Supplementary figures and tables for "Avidity as a marker of protection against hepatitis E during a genotype 1 outbreak in South Sudan"

**Supplementary Materials**

**Supplementary Figure S1. Flow diagram of patient enrollment and derivation of avidity-tested subset and analytic population**

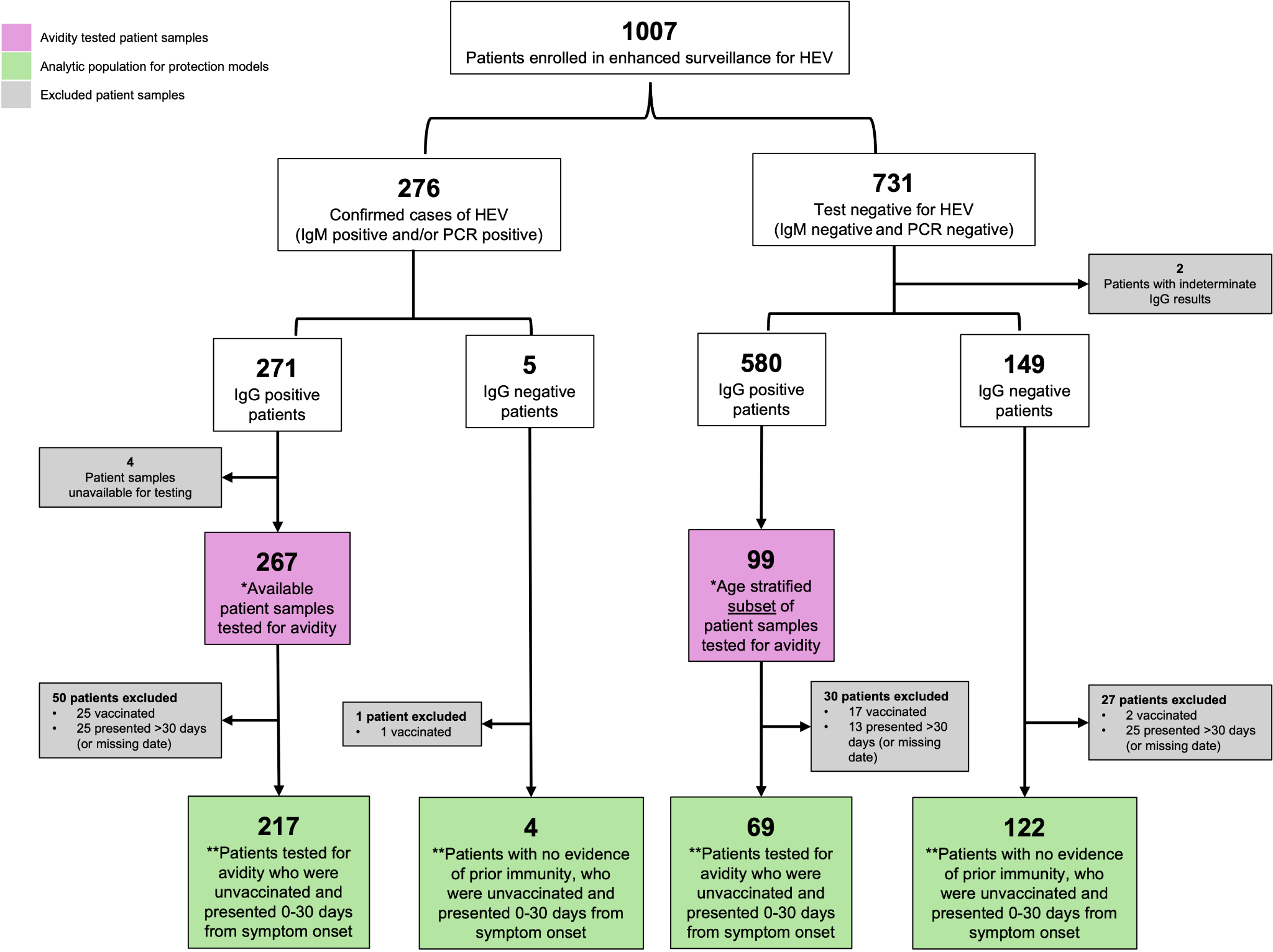

**Supplementary Figure S1: Flow diagram of patient enrollment and derivation of avidity-tested subset and analytic population.** A total of 1007 patients enrolled in enhanced surveillance in the Bentiu camp met the suspected HEV case definition for acute jaundice syndrome; 276 patients were confirmed cases of HEV (IgM-positive and/or PCR-positive) and 731 were test-negative (IgM- and PCR-negative).

*Avidity-tested subset (purple; n = 366): Anti-HEV IgG avidity was measured in all IgG-positive, HEV confirmed patients whose samples were available (267 of 271) and in an age-stratified subsample of IgG-positive test negative individuals (99 of 580; approximately 20 per age band: 0–5, 6–10, 11–15, 16–39, 40+ years). **Analytic population for protection models (green; n = 412): The 412 unvaccinated patients who presented within 30 days of symptoms onset and had an observed exposure (IgG-negative or IgG-positive individuals with measured avidity).

**Supplementary Figure S2: Avidity quality check**

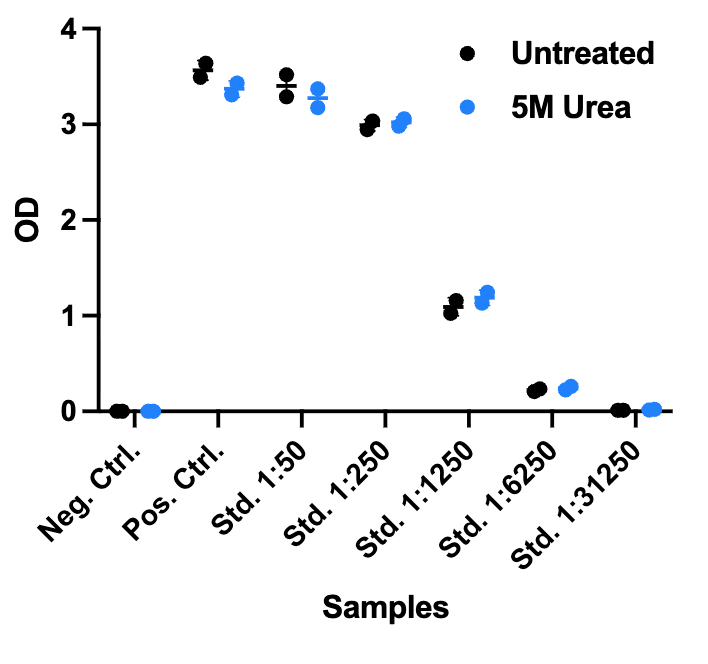

**Supplementary Figure S2: Avidity quality check**. Wantai IgG ELISA assays have been treated with 5M urea (blue dots) or wish buffer only (black dots) before addition of serum dilutions to address whether Urea treatment affects binding of antigen to the plate or antigen conformation. OD values are similar between treated and untreated samples indicating no effect of urea on the antigen.

**Supplementary Figure S3: Causal structure and identification**

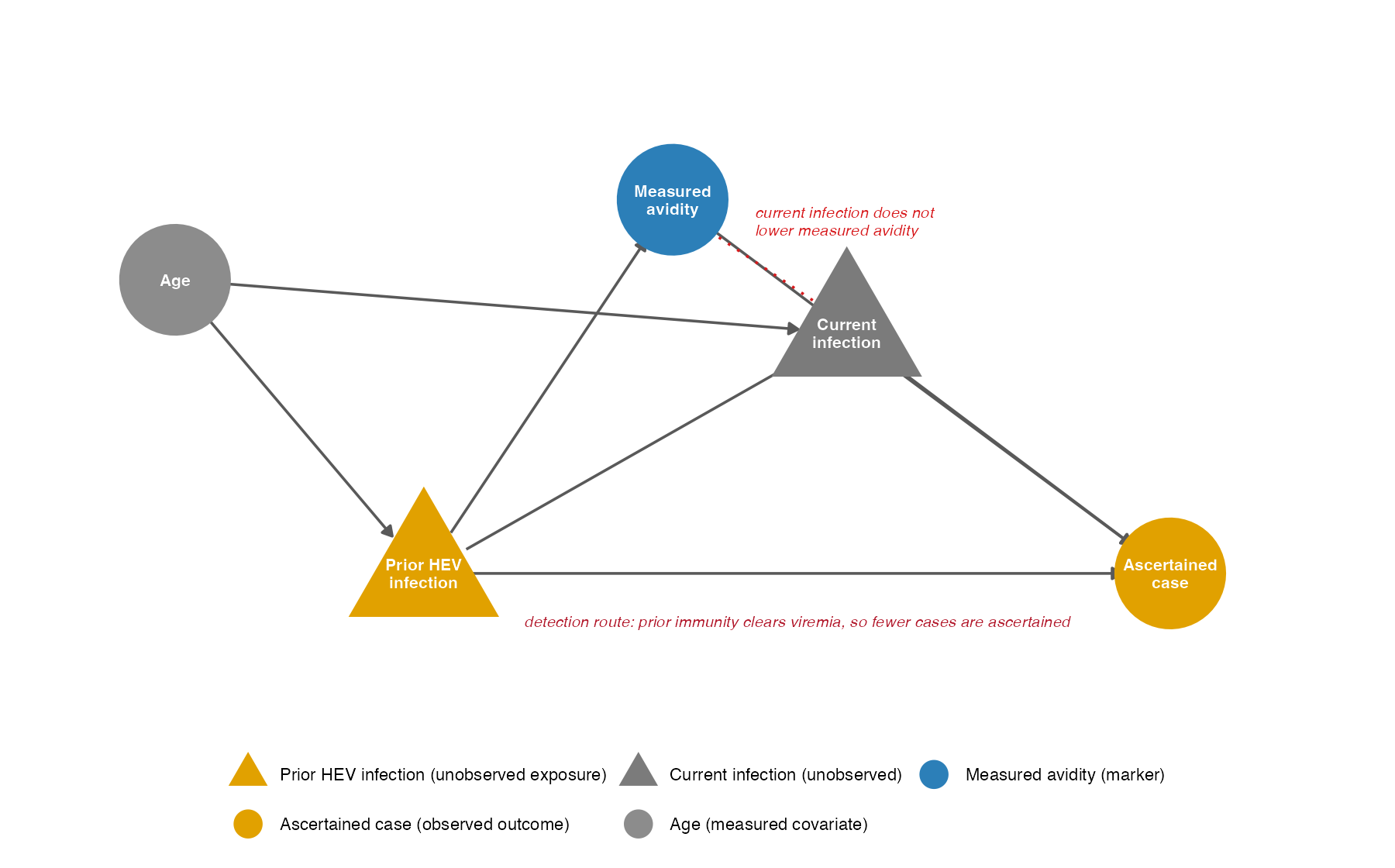

**Supplementary Figure S3: Causal structure and identification.** Triangular/grey nodes denote unobserved variables. Gold nodes are the estimand endpoints: prior HEV infection (exposure) and the ascertained case (observed outcome). Measured avidity (blue) is a specific but only moderately sensitive marker of prior infection; the current infection has no arrow into it (red, dotted, no arrowhead) because secondary antibody responses are memory-derived and of high avidity, so a current infection does not lower the measured avidity index (i.e. measured avidity is not a descendant of the outcome and exposure misclassification is non-differential). Prior immunity reaches the ascertained case by two routes that cannot be separated observationally: a protection route (prior → current infection → case) and a detection route (prior → case), the latter because immunity clears viremia so a true reinfection is less often PCR/IgM-detected. Age is the only measured common cause of prior and current infection; adjusting for it blocks all back-door paths.

**Supplementary Figure S4. Seroprevalence by age among test-negative participants**

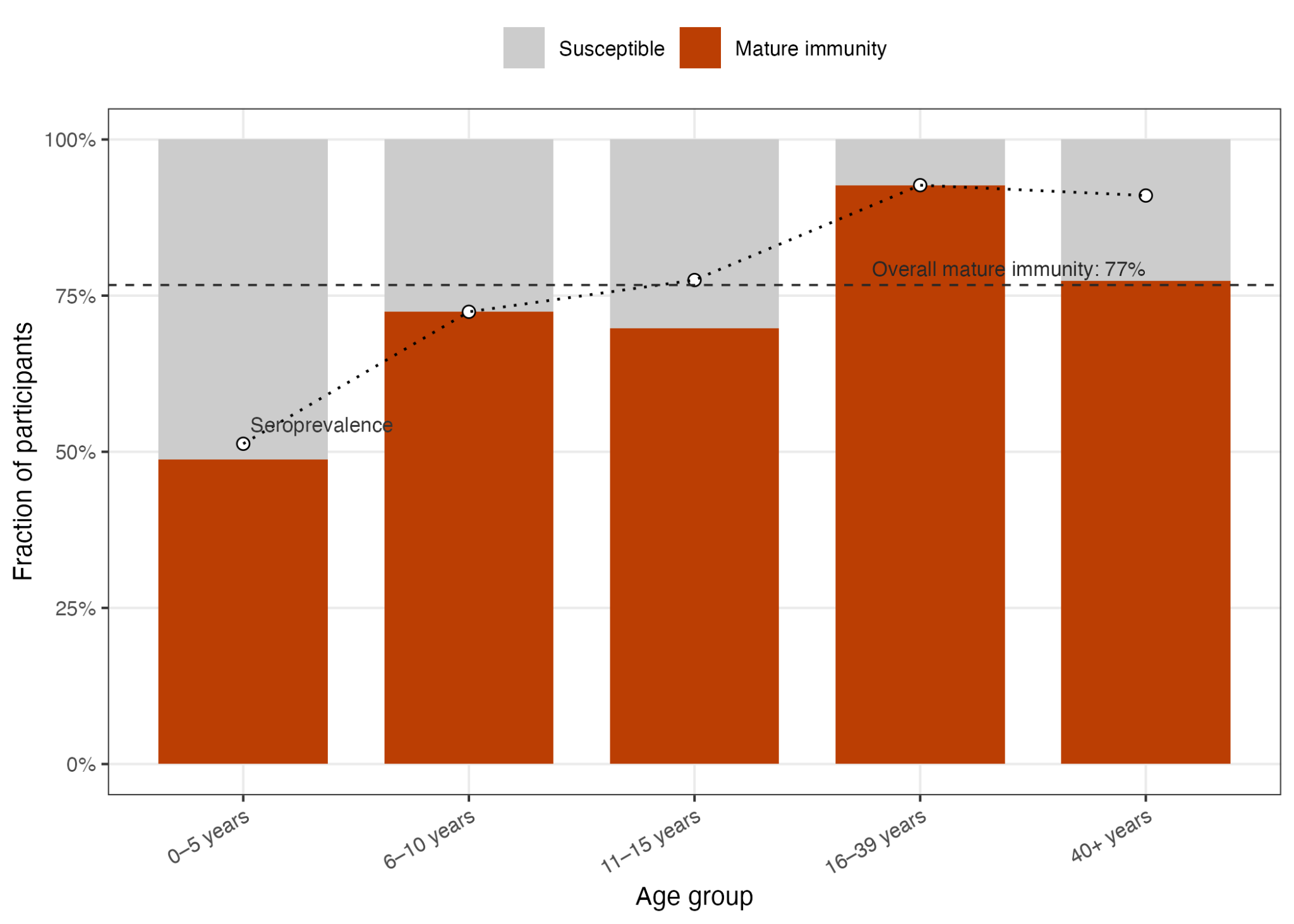

**Supplementary Figure S4: Seroprevalence by age among test-negative participants.** Mature immunity is weighted for the age-stratified sampling; the susceptible band is 1 - mature. Across all ages, an estimated 77% of test-negative participants had mature immunity.

**Supplementary Figure S5. Avidity maturation over time**

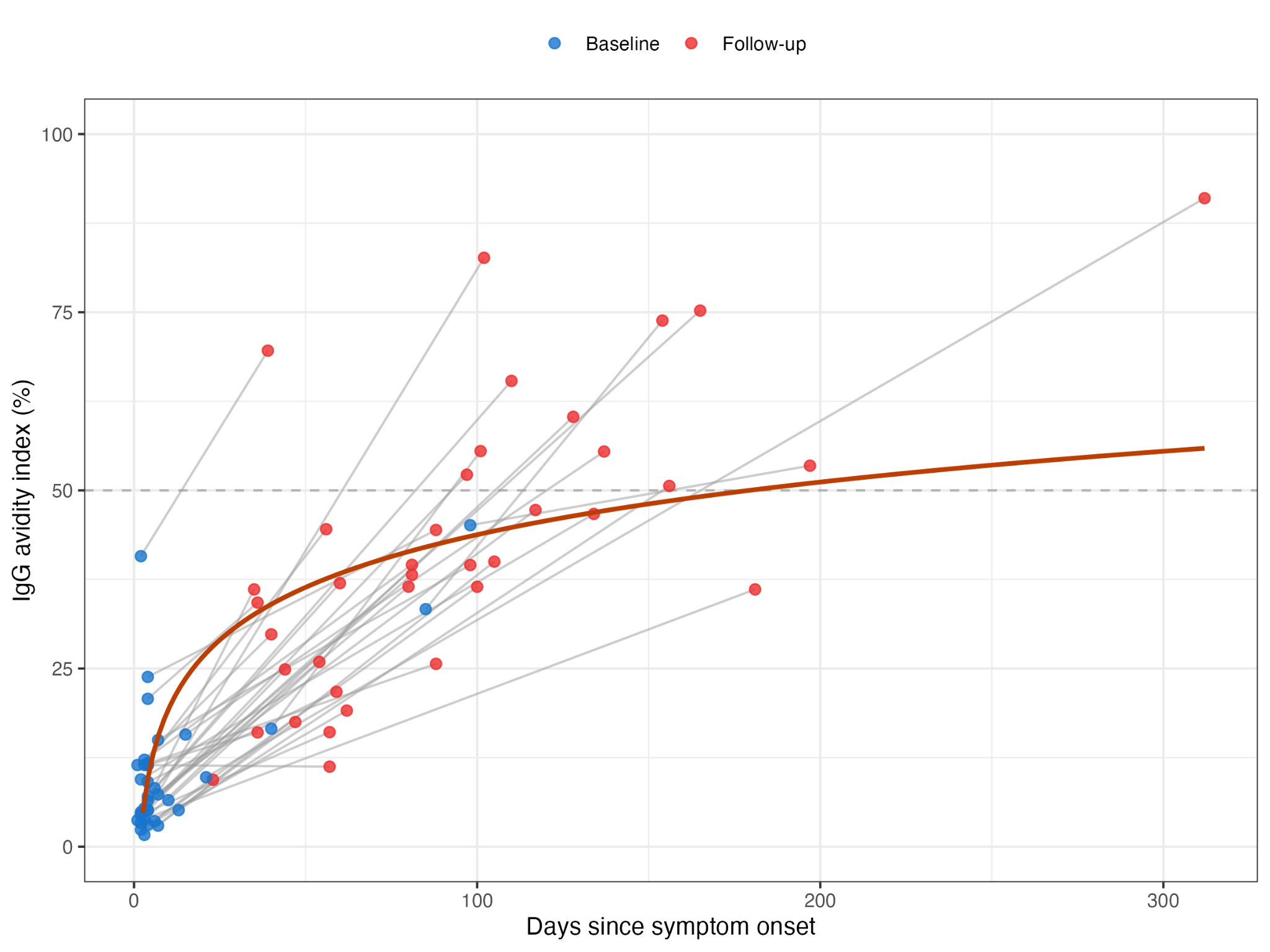

**Supplementary Figure S5: Avidity maturation over time.** Paired baseline and follow-up sera were available for 42 confirmed cases, but follow-up sampling spanned a wide window (median 84 days since onset, range 23–312), and follow-up avidity depended strongly on this timing (Spearman ρ = 0.52, p < 0.001). A single paired comparison therefore understates maturation. modeling avidity over time among primary infections (low baseline avidity; mixed model fit with lme4, avidity ~ log[days since onset], per-patient random intercept), avidity gradually increased with a predicted mean of 31% at 30 days, 43% at 90 days, and ~50% at ~180 days indicating that maturation to high avidity typically takes months. Accordingly, the fraction reaching high avidity by follow-up was almost entirely a function of when the follow-up occurred: 7% (1/14) among those sampled within 60 days of onset, 29% (4/14) at 61–120 days, and 78% (7/9) beyond 120 days. All five cases with high baseline avidity (reinfections) remained high.

**Supplementary Figure S6. Anti-HEV IgG levels (Luminex MFI) and viral load**

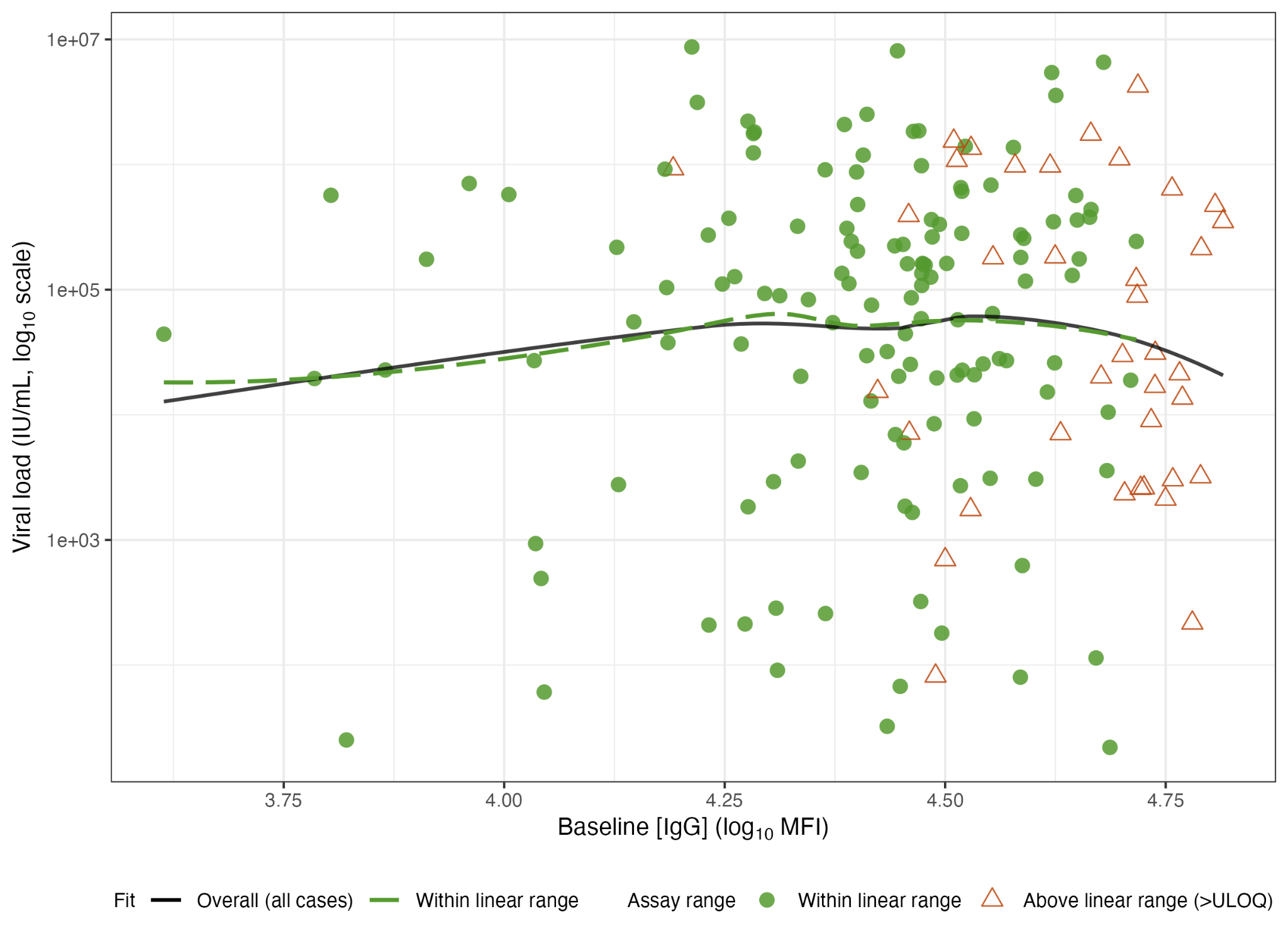
**Supplementary Figure S6: Anti-HEV IgG levels (Luminex MFI) and viral load.** Points represent unvaccinated confirmed HEV cases, with symbols and colors indicating where each sample fell relative to its plate’s standard-curve linear range: within range (green circles) or above the upper limit of quantification (>ULOQ; open orange triangles). No sample fell below the lower limit of quantification. Two LOESS fits summarize the relationship between baseline anti-HEV IgG levels (Luminex median fluorescence intensity (MFI)) and viral load: across all cases (black solid) and restricted to within-range cases (green-dashed). Both fits closely overlap and indicate no association between IgG and viral load.

**Supplementary Table S1: Sufficient adjustment sets by estimand**

| **Estimand** | **Sufficient adjustment set** | **Note** |
| --- | --- | --- |
| Prior infection → ascertained case (total effect) | {age} | Total effect on **detected** disease; conflates protection (prior→infection→case) with detection (prior→case, via viral clearance), which is not separable observationally. |
| Pre-infection avidity → ascertained case (direct) | none exists among observed | Not identified: the unobserved prior infection confounds it through non-avidity routes. |
| *Prior infection is unobserved; measured avidity is its marker. Measured avidity is not a descendant of the outcome (no current-infection → avidity edge), so exposure misclassification is non-differential. The avidity → case effect is not identified (the unobserved prior infection confounds it). The prior-infection → case effect is identified by {age}, but is a total effect on detected disease that conflates protection with detection. Sufficient adjustment sets verified by d-seperation in daggity.* | | |

**Supplementary Table S2: Characteristics of anti-HEV IgG positive study participants with available blood samples for avidity testing (n = 366)**

|  | **Confirmed-HEV case** | | **Test-negative** | |
| --- | --- | --- | --- | --- |
|  | **High (N=34)** | **Low (N=233)** | **High (N=93)** | **Low (N=6)** |
| **Age (years)** |  |  |  |  |
| Mean (SD) | 18.1 (14.1) | 12.0 (9.52) | 18.7 (17.2) | 37.8 (33.6) |
| Median [IQR] | 14.1 [6.95, 22.0] | 9.10 [4.60, 17.3] | 11.7 [5.90, 25.0] | 34.0 [10.6, 63.4] |
| **Sex** |  |  |  |  |
| Female | 11 (32.4%) | 110 (47.2%) | 45 (48.4%) | 2 (33.3%) |
| Male | 23 (67.6%) | 123 (52.8%) | 48 (51.6%) | 4 (66.7%) |
| **Days from symptom onset** |  |  |  |  |
| 0-5 days | 18 (52.9%) | 118 (50.6%) | 42 (45.2%) | 0 (0%) |
| 6-30 days | 12 (35.3%) | 92 (39.5%) | 39 (41.9%) | 2 (33.3%) |
| 31-100 days | 4 (11.8%) | 19 (8.2%) | 10 (10.8%) | 3 (50.0%) |
| Missing | 0 (0%) | 4 (1.7%) | 2 (2.2%) | 1 (16.7%) |
| **Effective vaccine doses** |  |  |  |  |
| 0 | 28 (82.4%) | 214 (91.8%) | 77 (82.8%) | 5 (83.3%) |
| 1 | 2 (5.9%) | 14 (6.0%) | 5 (5.4%) | 0 (0%) |
| ≥2 | 4 (11.8%) | 5 (2.1%) | 11 (11.8%) | 1 (16.7%) |
| **PCR result** |  |  |  |  |
| Negative | 24 (70.6%) | 51 (21.9%) | 93 (100%) | 6 (100%) |
| Positive | 10 (29.4%) | 182 (78.1%) | 0 (0%) | 0 (0%) |
| **Baseline IgM result** |  |  |  |  |
| Indeterminate | 1 (2.9%) | 1 (0.4%) | 3 (3.2%) | 0 (0%) |
| Negative | 7 (20.6%) | 3 (1.3%) | 90 (96.8%) | 6 (100%) |
| Positive | 26 (76.5%) | 229 (98.3%) | 0 (0%) | 0 (0%) |
| **Baseline anti-HEV (log_10_ MFI)** |  |  |  |  |
| Mean (SD) | 4.59 (0.249) | 4.47 (0.236) | 4.35 (0.499) | 3.46 (0.471) |
| Median [IQR] | 4.68 [4.54, 4.74] | 4.49 [4.37, 4.64] | 4.54 [4.12, 4.77] | 3.28 [3.26, 3.34] |
| **Baseline anti-HEV IgG avidity index (%)** |  |  |  |  |
| Mean (SD) | 74.8 (14.6) | 13.7 (11.7) | 78.4 (13.8) | 36.7 (18.0) |
| Median [IQR] | 73.0 [63.7, 88.9] | 9.77 [5.20, 16.9] | 79.1 [69.4, 88.2] | 43.1 [38.4, 46.3] |
| **ALT (U/L)** |  |  |  |  |
| Mean (SD) | 167 (319) | 460 (486) | 37.0 (70.7) | 34.5 (34.6) |
| Median [IQR] | 16.3 [11.2, 28.8] | 194 [16.8, 859] | 15.8 [12.2, 22.3] | 15.0 [14.5, 44.8] |
| Missing | 10 (29.4%) | 54 (23.2%) | 34 (36.6%) | 3 (50.0%) |
| **Total Bilirubin (µmol/L)** |  |  |  |  |
| Mean (SD) | 8.93 (28.0) | 18.5 (43.1) | 1.89 (3.47) | 3.25 (5.84) |
| Median [IQR] | 0.587 [0.500, 6.92] | 4.31 [0.500, 12.0] | 0.500 [0.500, 0.807] | 0.500 [0.375, 3.38] |
| Missing | 12 (35.3%) | 74 (31.8%) | 35 (37.6%) | 2 (33.3%) |
| *Characteristics of anti-HEV IgG positive study participants with available blood samples for avidity testing (N = 366), stratified by case status and IgG avidity category. Continuous variables are summarized as mean (SD) and median [min, max], and categorical variables as n (%). Vaccine doses were lagged by 14 days (counted as effective ≥14 days post-dose) using the same methods previously described [7].* | | | | |

**Supplementary Table S3: Protection from prior mature immunity against confirmed HEV disease, by model specification**

| **Model** | **aOR (95% CI)** | **Protection (95% CI)** | **p** |
| --- | --- | --- | --- |
| Weighted, ns(age,3) | 0.046 (0.025–0.084) | 95% (92–98%) | <0.001 |
| Unweighted, ns(age,3) | 0.189 (0.106–0.335) | 81% (66–89%) | <0.001 |
| Unweighted, linear age | 0.228 (0.133–0.392) | 77% (61–87%) | <0.001 |
| Weighted, PCR-confirmed cases only | 0.016 (0.006–0.040) | 98% (96–99%) | <0.001 |
| *Logistic regression in unvaccinated participants presenting within 30 days of onset. Exposure: mature IgG (avidity ≥50%) vs no evidence of prior immunity (IgG-negative, or IgG-positive with avidity <50%). Adjusted for age (natural cubic spline, 3 df, except the linear-age row), sex, and delay to care. The weighted model applies inverse-sampling weights for the subsampled IgG-positive test-negatives and is the primary estimate. All specifications are fit under a common survey design (svyglm), so 95% CIs are design-based Wald intervals throughout; the unweighted rows use unit weights. The final row is a case-definition sensitivity restricting cases to PCR-confirmed only (test-negatives unchanged): protection rises to ~98%, so the primary estimate is not driven by IgM-only case ascertainment.* | | | |

**Supplementary Table S4. Protection estimates across avidity thresholds (weighted, spline-adjusted)**

| **Avidity cutoff (%)** | **high-avidity n** | **Protection (95% CI)** |
| --- | --- | --- |
| 15 | 139 | 82% (71–89%) |
| 20 | 127 | 87% (78–92%) |
| 25 | 115 | 90% (84–94%) |
| 30 | 110 | 92% (86–95%) |
| 35 | 106 | 93% (88–96%) |
| 40 | 102 | 94% (89–96%) |
| 45 | 99 | 94% (90–97%) |
| 50 | 92 | 95% (92–98%) |
| 55 | 87 | 95% (90–97%) |
| 60 | 80 | 95% (90–98%) |
| 65 | 74 | 94% (88–97%) |
| 70 | 62 | 92% (83–96%) |
| 75 | 48 | 91% (81–96%) |
| 80 | 43 | 90% (77–95%) |
| 85 | 32 | 88% (69–95%) |
| 90 | 23 | 83% (54–94%) |
| 95 | 12 | 89% (45–98%) |

*Each row is a separate weighted, spline-adjusted model at a different avidity cutoff. The population is fixed; only the high/low classification moves, so the reference group changes at every step. Therefore, this illustrates threshold robustness, not a dose-response. In the weighted analysis, protection is high across every threshold (82-95% from a 15% to a 90% cutoff), peaking near the pre-specified 50% cutoff, so the finding does not depend on the threshold.*

**Supplementary Table S5. Competing jaundice causes among confirmed cases by infection status and avidity**

*A high-avidity "confirmed" case may not necessarily represent HEV-caused disease. In a previously immune person, an ascertained case can be ambiguous in two ways: an IgM-only case may reflect persisting or old IgM while the jaundice is another cause; and a PCR-positive case may reflect incidental low-level shedding - a trace of HEV RNA with no biochemical hepatitis - while the jaundice is another cause. Given that both routes would inflate the reinfection fraction and bias protection toward the null, we performed sensitivity analysis on the direction of the main findings. Among confirmed cases with a measured avidity, we recorded a competing cause of jaundice (positive malaria RDT, HBsAg, or HAV/HBV PCR) and found that high-avidity cases are about twice as likely to carry a competing cause, in both the IgM-only and PCR-positive strata.*

| **Infection status** | **Avidity** | **n** | **Malaria RDT+** | **HBsAg+** | **HAV/HBV PCR+** | **Any competing cause** | **Median log10 VL** | **Median ALT** |
| --- | --- | --- | --- | --- | --- | --- | --- | --- |
| IgM-only | Low | 51 | 5 (10%) | 4 (8%) | 0 (0%) | 9 (18%) | NA | 14 |
| IgM-only | High | 24 | 5 (21%) | 2 (8%) | 1 (4%) | 7 (29%) | NA | 12 |
| PCR-positive | Low | 182 | 19 (10%) | 10 (5%) | 6 (3%) | 32 (18%) | 5.1 | 500 |
| PCR-positive | High | 10 | 2 (20%) | 2 (20%) | 0 (0%) | 4 (40%) | 2.3 | 449 |
| *Any competing cause = positive malaria RDT, HBsAg, or HAV/HBV PCR. Any competing cause by avidity (all confirmed cases): Fisher OR 2.23 (95% CI 0.91–5.22), p = 0.061. Median viral load (log10 IU/mL) and ALT (U/L) among cases with a measurement.* | | | | | | | | |

**Supplementary Table S6. Protection (1 - adjusted OR) under two considerations of potential alternative cause cases**

*To test whether our original estimates could be an artifact of non-HEV jaundice being counted as HEV disease, we re-estimated protection accounting for potential alternative causes of jaundice. For each “potential-alt-cause” set and handling combination, we refit the primary model unchanged, redefining confirmed cases and test-negatives accordingly. Individuals in each set were classified in two ways, excluded (dropped) or treated as test-negative (reclassified as controls). Confirmed cases reclassified to the test-negative group were unweighted as they were not part of the IgG-positive control subsampling frame; all other test-negatives remained unchanged. We also refit a model with the case group restricted to PCR-confirmed cases (excluding IgM-only cases) as a separate case-definition check. Protection remained between 95% and 98% across all sets and handlings, indicating the estimate is not driven by potential competing causes and is robust to a stricter case definition.*

| **Potential-alt-cause set** | **Handling** | **Cases (n)** | **Protection (95% CI)** | **aOR (95% CI)** |
| --- | --- | --- | --- | --- |
| None (primary) | All confirmed cases | 221 | 95% (92–98%) | 0.046 (0.025–0.084) |
| Restrict to PCR-confirmed | IgM-only cases excluded | 166 | 98% (96–99%) | 0.016 (0.006–0.040) |
| Competing cause | Excluded | 177 | 97% (93–98%) | 0.033 (0.016–0.067) |
|  | Treated as test-negative | 177 | 96% (91–98%) | 0.044 (0.023–0.086) |
| Competing cause or shedding | Excluded | 175 | 97% (94–99%) | 0.031 (0.015–0.063) |
|  | Treated as test-negative | 175 | 96% (92–98%) | 0.042 (0.021–0.083) |
| *Weighted, spline-adjusted logistic model (unvaccinated, delay < 30 days), as in the primary analysis. 'Excluded' drops the cases; 'Treated as test-negative' reclassifies them as controls. Test-negatives are otherwise unchanged; reclassified controls enter unweighted (they were not part of the IgG-positive control subsampling frame). Restrict-to-PCR-confirmed shown for reference.* | | | | |
